# A Digital Survey on the Acceptance and Affordability of COVID 19 Vaccine among the People of West Bengal, India- A Survey Based Study

**DOI:** 10.1101/2020.11.13.20229534

**Authors:** Arunodaya Gautam, Bikram Dhara, Dattatreya Mukherjee, Debraj Mukhopadhyay, Sayan Roy, Soumya Sarathi Ganguly, Ankita Dutta Chowdhury, Shubham Goswami, Swapnil Dey, Srishti Basu, Deboshmita Banerjee, Soumi Chatterjee, Ishita Roy, Arup Kumar Mitra

## Abstract

**Overview:** Currently, multiple vaccines for coronavirus disease 2019 (COVID-19) are in clinical trials. In Oct-Nov 2020, 1078 individuals in West Bengal surveyed to evaluate possible acceptance rates, affordability and factors affecting the acceptance of a vaccine for COVID-19.

**Result:** 77.27 percent of respondents reported that they would be very or very likely to take a vaccine for COVID-19, 5.3 percent don’t want to take vaccine and 12.24 don’t know about their decision. In that 58 percent respondents want to take Indian Vaccine; 19 percent respondents want to take foreign vaccine. Other respondents can take any vaccine. The affordability, 40 percent respondents want a vaccine bellow 500 INR, 25 percent respondents want a vaccine of 500-1000 INR. 11 percent respondents want vaccine of over 1000 INR.

**Method of Study:** The google form is prepared with the questions on acceptance and affordability of vaccines. The form is circulated digitally among the people and then we have collected the data in excel. Based on the result we have prepared our statistical graphs.

**Conclusion:** Majority of Responders want a COVID 19 vaccine. Majority responders want Indian COVID19 Vaccine. Majority responders want a vaccine in a cost of below 500 INR.

## Introduction

It is expected that the COVID-19 pandemic will begin to inflict immense morbidity and mortality pressures thus seriously affecting populations and economies worldwide. Governments must be prepared to ensure access to and delivery of COVID-19 vaccines on a wide scale, in a equal manner, if and when safe and reliable vaccines become available. This would include ample capacity for the health sector, as well as strategies to increase trust in and understanding of the vaccine and those that administer it. In 2015, considering the availability of vaccination facilities, the World Health Organization (WHO) Strategic Advisory Committee of Immunization Experts described vaccine hesitancy as a ‘delay in approval or denial of vaccination’(1), which may differ in type and severity depending on when and where it occurs and what vaccine is involved, as has been verified in several studies (2,3). Vaccine hesitancy issues are rising globally (4); in particular, in 2019, WHO recognized it as one of the top ten global health risks. Vaccine hesitancy and disinformation in many countries poses major challenges to the achievement of coverage and population immunity (5,6). Governments, public health agencies and awareness organizations must be prepared to tackle hesitancy and improve vaccine awareness so that, where possible, the population supports immunization. Anti-vaccination groups are now lobbying against the need for a vaccine in several nations, with others questioning the presence of COVID-19 entirely. Misinformation sent across multiple networks may have a major effect on the approval of a COVID-19 vaccine. (7,8). Governments and communities must assess existing levels of readiness to obtain a COVID-19 vaccine that is potentially safe and successful and identify causes of hesitancy and or approval of the vaccine. Phizer COVID 19 vaccine is 90% effective in SARS-COV2 infection COVID19 and it will be distributed from dec.(9) We are surveying on 1078 respondents in the focus of willingness and affordability of the vaccine.

## Methodology of the study

This is a survey based study which is done in west Bengal, a state in India. The Capital of West Bengal is India. In this survey a questionnaire is prepared on the acceptance and the affordability of COVID 19 vaccine. The questionnaire is prepared in google form and the google form is circulated among the people of West Bengal. The Data is taken in Excel sheet and the statistical graph is prepared in SSPS.

The Questionnaire is

|  |  |
| --- | --- |
| Consent form: This is a Survey based study where we want to know that you want to take vaccine or not when it will be available in market. Your all details will be confidential. This will take 2 minutes only. Thank You | YES/NO |
| Age group | 15-25/25-35/35-45/45-55/55-65/65+ |
| Educational qualification | Upto 10 or below/ Upto 12/ Graduation/Post Graduation/ Doctoral/ Post Doctoral |
| Gender | Male/Female/Others |
| Do you want to take COVID 19 vaccine when it will be commercially available? | Yes/No/Don't know |
| If No then why don't want:? |  |
| If Yes, then You want vaccine of | Indian Vaccine/Foreign Vaccine/Any Vaccine |
| Your affordable cost of vaccine | below 500/below1000/<br>above 1000 |

## Result

We have collected the data of 1078 people from West Bengal. Most of the people.

### The acceptance rate of Vaccine: (n=1078)

| Want to take vaccine | Don't want to take vaccine | Still Not Decided |
| --- | --- | --- |
| 77.27% (833) | 5.3% (58) | 12.2% (132) |

55 (5.1%) Respondents place this blank as they don't want to convey any decision

**Fig1:**
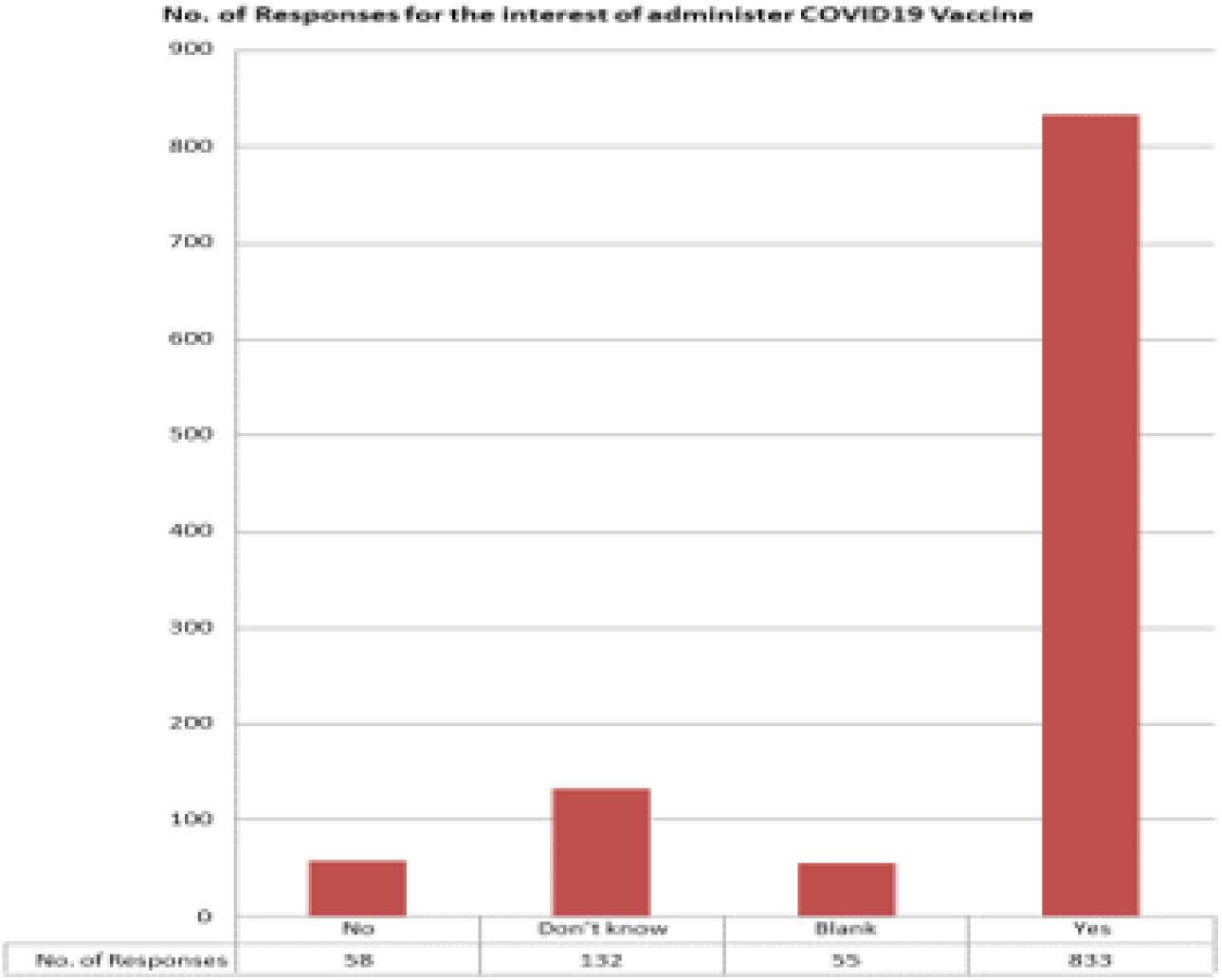
The acceptance rate of COVID 19 Vaccine.

### Indian Vaccine or Foreign Vaccine, Preference upon the manufacturing of the vaccine (n=1078)

Majority of respondents want an Indian vaccine. Although many people want a vaccine which will be available first in the market.

| Indian Vaccine | Foreign Vaccine | Any Vaccine |
| --- | --- | --- |
| 58% | 19% | 23% |

**Fig 2.1, 2.2:**
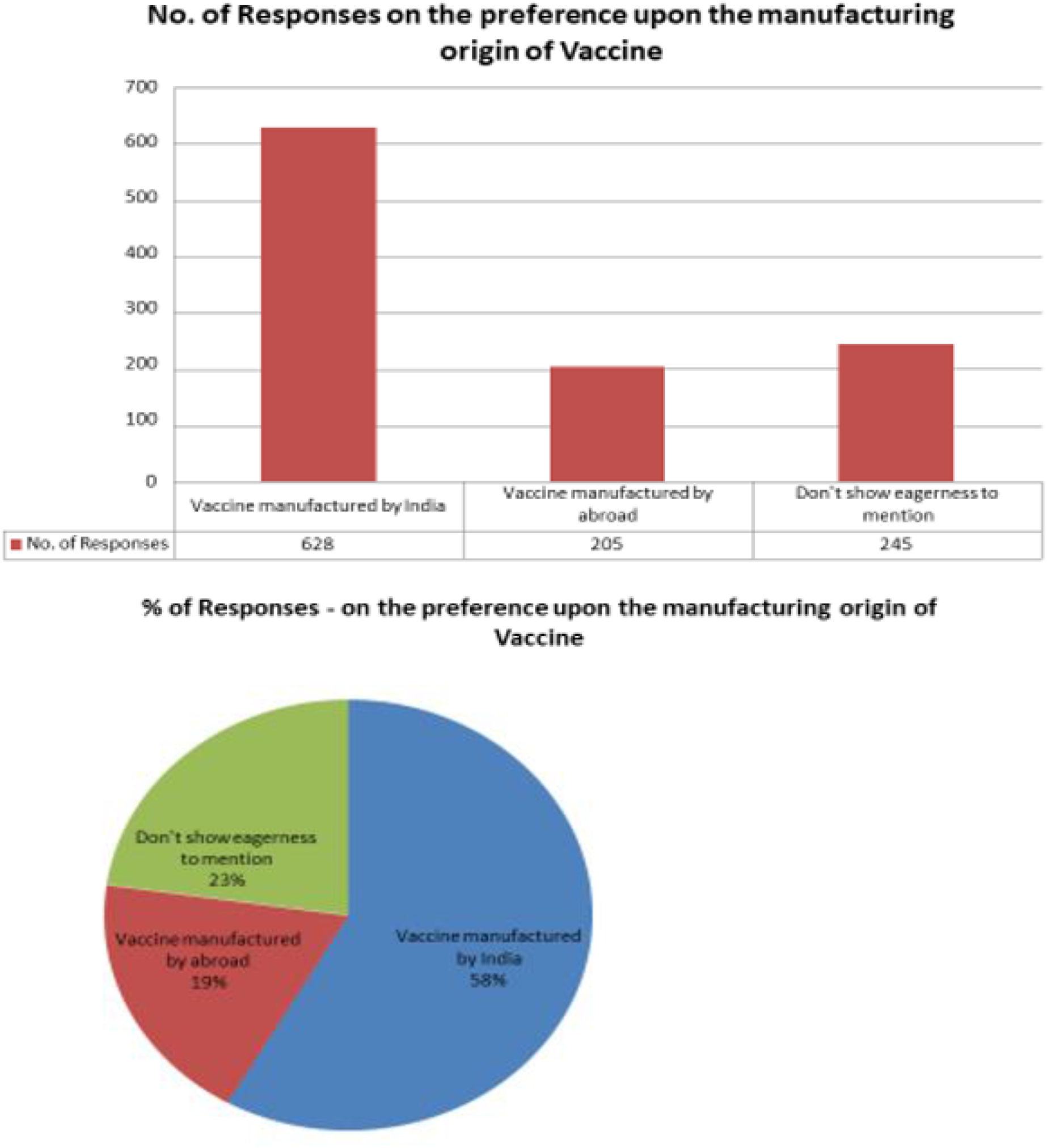
Preference upon the manufacturing origin of the vaccine.

### Affordability of the Vaccine: (n=1078)

Most of the responders want a cheap Vaccine which costs below 500 INR.

|  |  |  |  |
| --- | --- | --- | --- |
| Below 500 INR | 500-1000 INR | >1000 INR | Don't want to mention |
| 40% | 25% | 12% | 23% |

**Fig 3.1,3.2:**
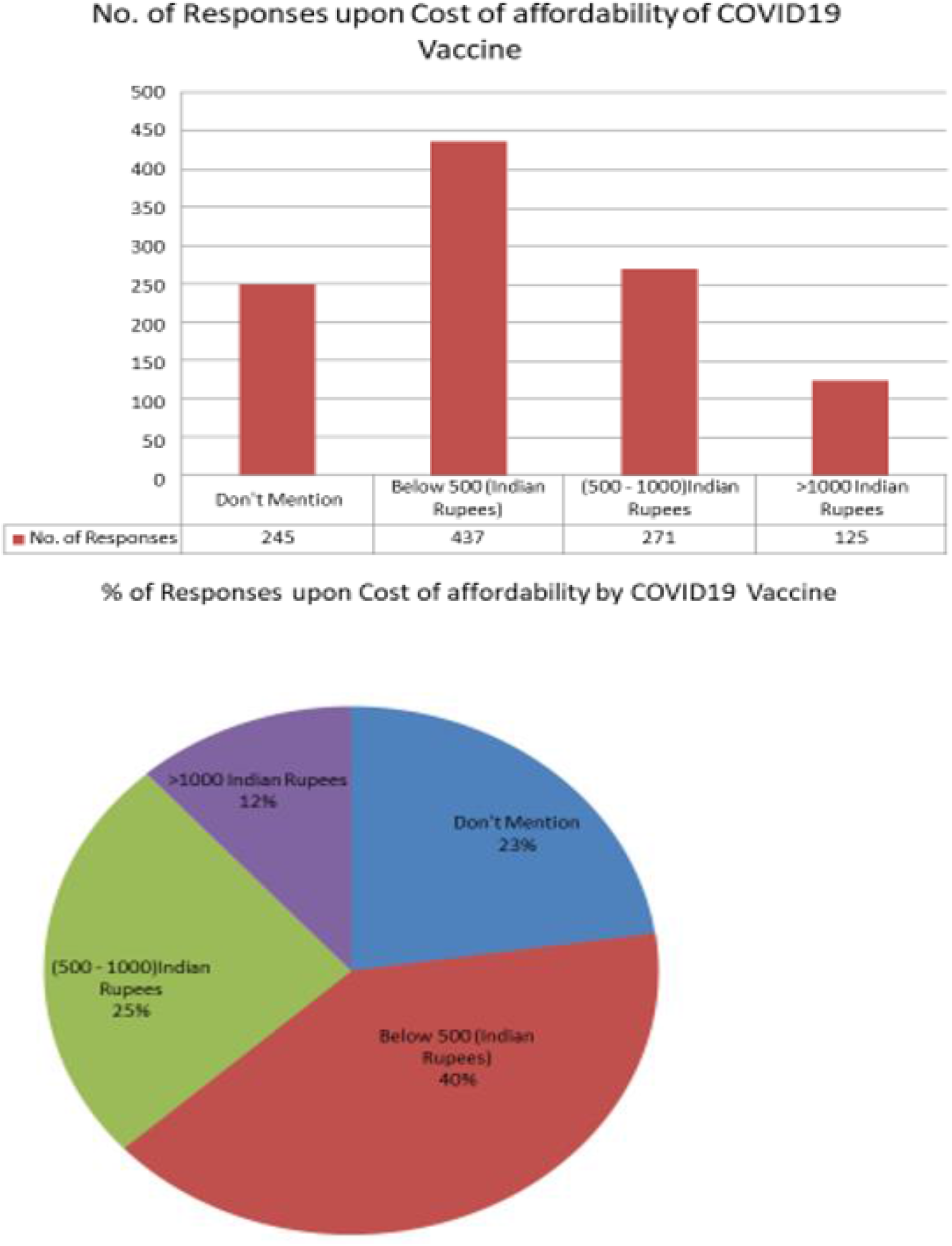
Affordability of COVID19 vaccine.

## Discussion

This study is based on a Spanish study which is published in Nature.(10) Our study is focused in only one state in India that is West Bengal. We have seen that most of the responders eagerly waiting for a vaccine which is a good sign. Although some responders have many misconceptions and educational gaps for which they don’t want to take a vaccine. A proper awareness is needed to them. Responders will prefer a Indian Vaccine although they have informed that which vaccine will come first in the market they will take irrespective of any origin. The responder’s affordability is checked in this study where most of the responders wants a cheap vaccine or a free vaccine from the government.

This study shows people eagerly waiting for a COVID19 vaccine. Although there are some limitation in the study. We made it digital survey, so the class of responders have a basic education and have a smart phone. Due to COVID 19 situation we are unable to reach the slam area community people. Some people don’t know anything about vaccine so there is an educational gap. A proper awareness about vaccination is needed.

## Conclusion

Majority of Responders want a COVID 19 vaccine. Majority responders want Indian COVID19 Vaccine. Majority responders want a vaccine in a cost of below 500 INR.

## Data Availability

Data are collected from the survey among the 1078 people of west bengal

## Author’s Contributions

Project Guide: Dr AK Mitra

Conceptualization of Project: D Mukherjee

Project Heads: A. Gautam, B Dhara, D Mukherjee

Protocol Preparation: All the Authors

Google Form Preparation: S. Roy

Survey: All the Authors

Statistics: D Mukhopadhyay, A Gautam

Paper Writing and Review: B Dhara, D Mukhopadhyay, D Mukherjee, S Chatterjee

Proof reading: A Gautam, A Dutta Chowdhury, S Chatterjee, SS Ganguly

## REFERENCES

1. MacDonald, N. E. & SAGE Working Group on Vaccine Hesitancy. Vaccine hesitancy: definition, scope and determinants. Vaccine 33, 4161–4164 (2015).

2. Karafillakis, E., Larson, H. J. & ADVANCE Consortium. The benefit of the doubt or doubts over benefits? A systematic literature review of perceived risks of vaccines in European populations. Vaccine 35, 4840–4850 (2017).

3. Cobos Muñoz, D., Monzón Llamas, L. & Bosch-Capblanch, X. Exposing concerns about vaccination in low- and middle-income countries: a systematic review. Int. J. Public Health 60, 767–780 (2015).

4. European Parliament. European Parliament resolution of 19 April 2018 on vaccine hesitancy and drop in vaccination rates in Europe (2017/2951 RSP). https://www.europarl.europa.eu/doceo/document/TA-8-2018-0188_EN.pdf (2018).

5. Larson, H. J., Jarrett, C., Eckersberger, E., Smith, D. M. D. & Paterson, P. Understanding vaccine hesitancy around vaccines and vaccination from a global perspective: a systematic review of published literature, 2007-2012. Vaccine 32, 2150–2159 (2014).

6. Lane, S., MacDonald, N. E., Marti, M. & Dumolard, L. Vaccine hesitancy around the globe: analysis of three years of WHO/UNICEF Joint Reporting Form data—2015– 2017. Vaccine 36, 3861–3867 (2018).

7. Enserink, M. & Cohen, J. Fact-checking Judy Mikovits, the controversial virologist attacking Anthony Fauci in a viral conspiracy video. Science https://www.sciencemag.org/news/2020/05/fact-checking-judy-mikovits-controversial-virologist-attacking-anthony-fauci-viral (2020).

8. Cornwall, W. Officials gird for a war on vaccine misinformation. Science 369, 14–19 (2020).

9. https://www.ndtv.com/world-news/us-plans-to-distribute-pfizer-covid-19-vaccine-in-december-health-secretary-alex-azar-2323589

10. Lazarus, J.V., Ratzan, S.C., Palayew, A. et al. A global survey of potential acceptance of a COVID-19 vaccine. Nat Med (2020). https://doi.org/10.1038/s41591-020-1124-9

